# Pericarditis and myocarditis long after SARS-CoV-2 infection: a cross-sectional descriptive study in health-care workers

**DOI:** 10.1101/2020.07.12.20151316

**Authors:** Rocio Eiros, Manuel Barreiro-Perez, Ana Martin-Garcia, Julia Almeida, Eduardo Villacorta, Alba Perez-Pons, Soraya Merchan, Alba Torres-Valle, Clara Sánchez Pablo, David González-Calle, Oihane Perez-Escurza, Inés Toranzo, Elena Díaz-Pelaez, Blanca Fuentes-Herrero, Laura Macías-Alvarez, Guillermo Oliva-Ariza, Quentin Lecrevisse, Rafael Fluxa, Jose L Bravo-Grande, Alberto Orfao, Pedro L Sanchez

## Abstract

**Background:** Cardiac sequelae of past SARS-CoV-2 infection are still poorly documented. We conducted a cross-sectional study in health-care workers to report evidence of pericarditis and myocarditis after SARS-CoV-2 infection.

**Methods:** We studied 139 health-care workers with confirmed past SARS-CoV-2 infection (103 diagnosed by RT-PCR and 36 by serology). Participants underwent clinical assessment, electrocardiography, laboratory tests including immune cell profiling and cardiac magnetic resonance (CMR). Pericarditis was diagnosed when classical criteria were present, and the diagnosis of myocarditis was based on the updated CMR Lake-Louise-Criteria.

**Results:** Median age was 52 years (IQR 41–57), 100 (72%) were women, and 23 (16%) were previously hospitalized for Covid-19 pneumonia. At examination (10.4 [9.3–11.0] weeks after infection-like symptoms), all participants presented hemodynamic stability. Chest pain, dyspnoea or palpitations were observed in 58 (42%) participants; electrocardiographic abnormalities in 69 (50%); NT-pro-BNP was elevated in 11 (8%); troponin in 1 (1%); and CMR abnormalities in 104 (75%). Isolated pericarditis was diagnosed in 4 (3%) participants, myopericarditis in 15 (11%) and isolated myocarditis in 36 (26%). Participants diagnosed by RT-PCR were more likely to still present symptoms than participants diagnosed by serology (73 [71%] vs 18 [50%]; p=0.027); nonetheless, the prevalence of pericarditis or myocarditis was high in both groups (44 [43%] vs 11 [31%]; p=0.238). Most participants (101 [73%]) showed altered immune cell counts in blood, particularly decreased eosinophil (37 [27%]; p<0.001) and increased CD4^-^CD8^-/lo^Tαβ-cell numbers (24 [17%]; p<0.001). Pericarditis was associated with elevated CD4^-^CD8^-/lo^Tαβ-cell numbers (p=0.011), while participants diagnosed with myopericarditis or myocarditis had lower (p<0.05) plasmacytoid dendritic cell, NK-cell and plasma cell counts and lower anti-SARS-CoV-2-IgG antibody levels (p=0.027).

**Conclusions:** Pericarditis and myocarditis with clinical stability are frequent long after SARS-CoV-2 infection, even in presently asymptomatic subjects. These observations will probably apply to the general population infected and may indicate that cardiac sequelae might occur late in association with an altered (delayed) innate and adaptative immune response.

The trial is registered with ClinicalTrials.gov, number NCT04413071

**Research in context:** *Evidence before this study:* Very little evidence exists describing long cardiac sequelae after SARS-CoV-2 infection. Although pericarditis and myocarditis are the two most frequent cardiac manifestations observed after a viral infection, as of May 13, 2020, the peer-reviewed literature was limited to isolated case reports of myocarditis and pericarditis during the COVID-19 hospitalization phase and to a retrospective observation in 26 recovered patients with COVID-19 pneumonia presenting cardiac complaints during hospitalization, revealing the presence of myocardial oedema in 14 (54%) patients and late gadolinium enhancement in 8 (31%) patients. These small size case series, limited to hospitalized RT-PCR patients with COVID-19 pneumonia, are insufficient to generalize conclusions about the true prevalence of pericardial and myocardial long involvement after SARS-CoV-2 infection. In addition, no study has investigated the immunological consequences of SARS-CoV-2 infection in the settings of pericarditis and myocarditis.

*Added value of this study:* To our knowledge, this is the largest cohort of subjects (N=139) —even for other common viruses— with clinical, electrocardiographic, laboratory and CMR imaging evaluations, to assess pericardial and myocardial involvements after SARS-CoV-2 infection. The strength of this study is the addition of non-hospitalized participants and also the inclusion of participants diagnosed of past SARS-CoV-2 infection through serology. Contrary to previous studies, women are well represented. We found a prevalence of pericarditis or myocarditis up to 40% cases; pericarditis coexisted with some degree of concurrent myocardial inflammation in 11% cases. Study participants who were previously hospitalized for COVID-19 pneumonia and patients who received antiviral (hydroxychloroquine, lopinavir-ritonavir) or anti-inflammatory (high-dose glucocorticoids and anti-interleukin treatments) treatments, and who were on chronic drug treatment with statins, were less likely to develop pericarditis or myocarditis. The clinical assessment of the participants showed clinical stability without any patient presenting severe pericardial effusion, heart failure or left ventricular dysfunction. We provide new data on seropositive subjects; although RT-PCR participants were more likely to still present symptoms than participants diagnosed by serology, the prevalence of pericarditis, myocarditis or myocarditis, almost three months after the initial viral prodrome, was high in both groups. In-depth investigation of the distribution of multiple major and minor populations of immune cells in blood showed high frequency of altered immune profiles after SARS-CoV-2 infection. The altered immune cell profiles identified partially mimic abnormalities previously reported during active infection together with others described here for the first time, with unique patterns associated with pericardial and/or myocardial injury. Nonetheless, we also described altered immune profiles in participants without pericardial and myocardial manifestations. Whether these later alterations are due to persistence of tissue damage in other organs affected by SARS-CoV-2, such as the lung, or they reflect normal post-infection immune recovery mechanisms, remains to be investigated.

*Implications of all the available evidence:* At present, there is much interest in the long-term sequelae of COVID-19. It is intriguing that pericarditis and myocarditis were observed so long after SARS-CoV-2 infection and also in some presently asymptomatic subjects, in association with notably altered immune cell profiles in blood. These observations will probably apply to the general population infected and may indicate that cardiac sequelae might occur late, paving the way for a better understanding the immune mechanisms involved. Thus, our study may have health-care consequences given the widespread diagnosis of SARS-CoV-2 infection in population-based seroprevalence studies.

## Introduction

Pericarditis and myocarditis are the two most frequent cardiac manifestations observed after a viral infection^1,2^. Symptoms tend to be non-specific and most cases resolve without long-term sequelae. That is why the true incidence of pericarditis and myocarditis after common viral infections —influenza, parvovirus, coxsackievirus, echovirus, adenovirus, echovirus, herpesvirus or cytomegalovirus— is still unknown in the general population.

The novel severe acute respiratory syndrome-coronavirus-2 (SARS-CoV-2) is currently causing a sustained covid-19 pandemic, with the risk of causing long-term cardiac sequelae in the infected population^3^. The fear of SARS-CoV-2 causing greater myocardial damage than other conventional viruses is based on its mechanism of infecting human cells by binding to the transmembrane angiotensin-converting-enzyme-2^4^, which is mainly expressed by cells in alveoli and myocardial tissue^5^; the rise in troponin levels observed in covid-19 patients hospitalized with pneumonia and its association with increased mortality^6,7^; and the probably reduced innate antiviral defences against a novel virus^8^.

Pericarditis and myocarditis after conventional viral infections both stem from an inadequate or excessive immune response driven by T and B cell-mediated mechanisms^1,2,9^. In case of an inadequate response, continued viral replication in the peri-myocardium protracts inflammation by attracting killer T cells and the concomitant production of chemokines and cytokines. In contrast, molecular mimicry can result in the production of autoantibodies against cardiac proteins, leading to a cardio-specific autoimmune response that causes sustained inflammation, effusion or cardiac remodeling. However, the specific immune profiles that occur after SARS-CoV-2 infection, particularly in patients presenting with cardiac sequelae remain unknown^10^.

The present study was designed to search for evidence of pericardial and myocardial involvement after past SARS-CoV-2 infection comprehensively studied by clinical assessment, laboratory tests, electrocardiography and cardiac magnetic resonance (CMR) imaging. Additionally, participants underwent an in-depth characterization of the immune cell compartments in blood and the virus-specific humoral immune response in this clinical scenario. As health-care workers have been the group most affected by SARS-CoV-2 in Spain, but have also been subject to more testing than the rest of the population, we decided to conduct the study in this singular cohort.

## Methods

### Study design and health-care workers participants

This cross-sectional, observational, cohort study consecutively recruited 142 health-care workers with laboratory confirmed SARS-CoV-2 infection in Salamanca, Spain, and who volunteered for the study. Among them, 106 health-care workers tested positive for SARS-CoV-2 by RT-PCR between March 13 and April 25; and 36 health-care workers were diagnosed after testing positive for anti-SARS-CoV-2-IgG antibodies between April 10 and May 22. The purpose of this second group was to also provide data from subjects with past SARS-CoV-2 infection in whom symptoms of viral infection are more likely to be mild and because population-based SARS-CoV-2 seroprevalence studies are becoming more established^11,12^. Study enrolment began on May 25 and finished on June 12, 2020.

Institutional approval (2020/05/490) for the study was provided by the University Hospital of Salamanca Ethics Committee, and all participants provided written informed consent. The study is registered with ClinicalTrials.gov NCT04413071. The responsibility for the study design, data collection and data interpretation lay solely with the study investigators. An internal adjudication monitoring board reviewed all cardiac study findings and adjudicated study outcomes. The authors had full access to all the data and elaborated all materials to submit for publication.

### Investigation process and procedures

All participants underwent clinical evaluation, electrocardiography, laboratory tests and CMR imaging at the same visit. After obtaining written informed consent, trained interviewers used a structure questionnaire to collect baseline data in face-to-face interviews. A cardiologist took a complete medical history, performed a physical examination and reviewed the completeness of the questionnaire in a separate room, where an electrocardiogram was performed, and blood samples were drawn immediately before the CMR. Electrocardiograms were interpreted in consensus by two experienced readers, who were blinded to participant identification, clinical history, symptoms, physical examination and other findings.

CMR was performed using a clinical 1.5 whole-body magnetic resonance scanner in the cardiac imaging laboratory of the University Hospital of Salamanca^13^. Functional imaging was performed using standard segmented cine steady-state free-precession sequence with breath holding. Myocardial oedema was identified using a T2-weighted short-tau triple inversion-recovery (T2W-STIR) sequence. T2-relaxation time properties were obtained using a T2-gradient spin-echo (T2-GraSE) mapping sequence. Assessment of T1-relaxation times and free water content within the myocardium (extracellular volume) were identified using a modified look-locker inversion recovery-MOLLI-with-5(3)3 acquisition scheme before and 15 minutes after intravenous administration of 0.15 mmol per kg body weight of gadobutrol contrast media agent. Late gadolinium enhancement was identified on a series of T1-weighted inversion recovery turbo field echo sequence, acquired 10 to 15 minutes after the contrast administration. CMR images were globally and regionally analysed using dedicated software, in consensus by two experienced readers, who were blinded in a similar manner to the electrocardiogram protocol, following conventional CMR methods (supplementary methods). T2 and T1-based markers of myocardial inflammation were analysed in each of the 16 segments of the 17-segment model of the American Heart Association (the true apex was excluded)^14^, where only positive segment concordances from the different T2 and T1-based markers were considered. Because myocarditis was diagnosed according to these T2 and T1-based CMR markers and an adequate selection of normal reference values is fundamental, we used as controls CMR imaging from 20-sex-and-aged matched individuals without cardiac disease from the general population of the province of Salamanca, Spain (NCT03429452)^15^.

Immunophenotypic analysis of (>250) immune cell populations was performed in peripheral blood samples collected in K3-EDTA (10mL/sample) and stained with the EuroFlow lymphocyte screening tube (LST) and the TCD4, NK/TCD8, BIgH and MoDC immune monitoring (IMM) tubes by flow cytometry (FACSCANTO II and LSR-Fortessa, respectively; Becton/Dickinson Biosciences, San José, California) using a dual-platform assay previously described in detail^16,17^. All protocols are available at www.euroflow.org. Reference values were defined based on a cohort of 463 age-matched adults (median age 52 years [IQR 47-61]) from the general population of the province of Salamanca, Spain. Anti-SARS-CoV-2-IgM (AnshLabs, Webster, Texas), IgG and IgA (Mikrogen Diagnostik, Neuried, Germany) antibody levels were measured in parallel in plasma from the same blood samples using commercially available IVD approved (semi-quantitative) ELISA kits, strictly as instructed by the manufacturers.

### Study outcomes and definitions

Study outcome measures were the prevalence of pericarditis and of myocarditis. Pericarditis was diagnosed if at least two of the following criteria were present, following current guidelines^1^: pericarditic chest pain, pericardial rub on auscultation, widespread ST-elevation or PR depression on ECG, and evidence of pericardial effusion at CMR. Elevation of inflammation markers, C-reactive protein, and evidence of pericardial inflammation at CMR were used as additional supporting findings. The diagnosis of myocarditis was based on the updated CMR Lake-Louis-Criteria (LLC)^18^, which consider as main LLC criteria for myocarditis positive oedema-sensitive T2-based markers (T2-weighted images or T2-mapping) or positive T1-based tissue characterization markers (abnormal T1-relaxation time or extracellular volume or late gadolinium enhancement), and as supportive LLC criteria either pericardial effusion, or evidence of pericardial inflammation at CMR, or systolic left ventricle wall motion abnormalities. Considering that participants were being examined beyond the acute phase of SARS-CoV-2 infection; myocarditis was defined as having a combination of at least two T2 or T1-based LLC main criteria or having a combination of only one T2 or T1-based LLC main criterion with one additional LLC supportive criterion.

As we were aware that pericarditis and myocarditis occur together in clinical practice, we hence defined as myopericarditis those cases of pericarditis with associated myocarditis on CMR but without left ventricle wall motion abnormalities, and as perimyocarditis those cases where left ventricle wall motion abnormalities were present^19^.

### Statistical Analysis

Descriptive statistics were used to summarized the data; results are presented as the proportion (%) of valid cases for categorical variables and as the median (IQR) for continuous variables. As the participants in our study were not randomly selected, all statistics are deemed descriptive only; nonetheless, differences between groups are also provided and were analysed by Fisher’s exact test for categorical variables and by nonparametric Mann-Whitney or Kruskal-Wallis for continuous data. We compared characteristics of participants and examinations, all Tables, according to the final clinical diagnosis (non-pericardial and myocardial manifestations vs pericarditis vs myopericarditis vs myocarditis). For 2-dimensional visualization of flow cytometry data, multivariate canonical analysis with multidimensional reduction of data via linear discriminant analysis, and the t-distributed stochastic neighbour embedding (t-SNE) machine-learning algorithm visualization tools, were used (Infinicyt software, Cytognos, Salamanca, Spain)^20^.

## Results

### Study Population

**Figure 1** depicts the flowchart for participant selection from the health-care workers. From the 142 recruited health-care workers who signed informed consent, one participant did not complete the CMR for claustrophobia. Two additional participants were excluded because history of severe hypertrophic myocardiopathy in one case, and inherited immune deficiency in the other. Thus, a total of 139 participants completed clinical assessment, electrocardiography, laboratory tests and CMR. Of these, 103 (74%) had been diagnosed by RT-PCR and 36 (26%) by serology.

**Figure 1:**
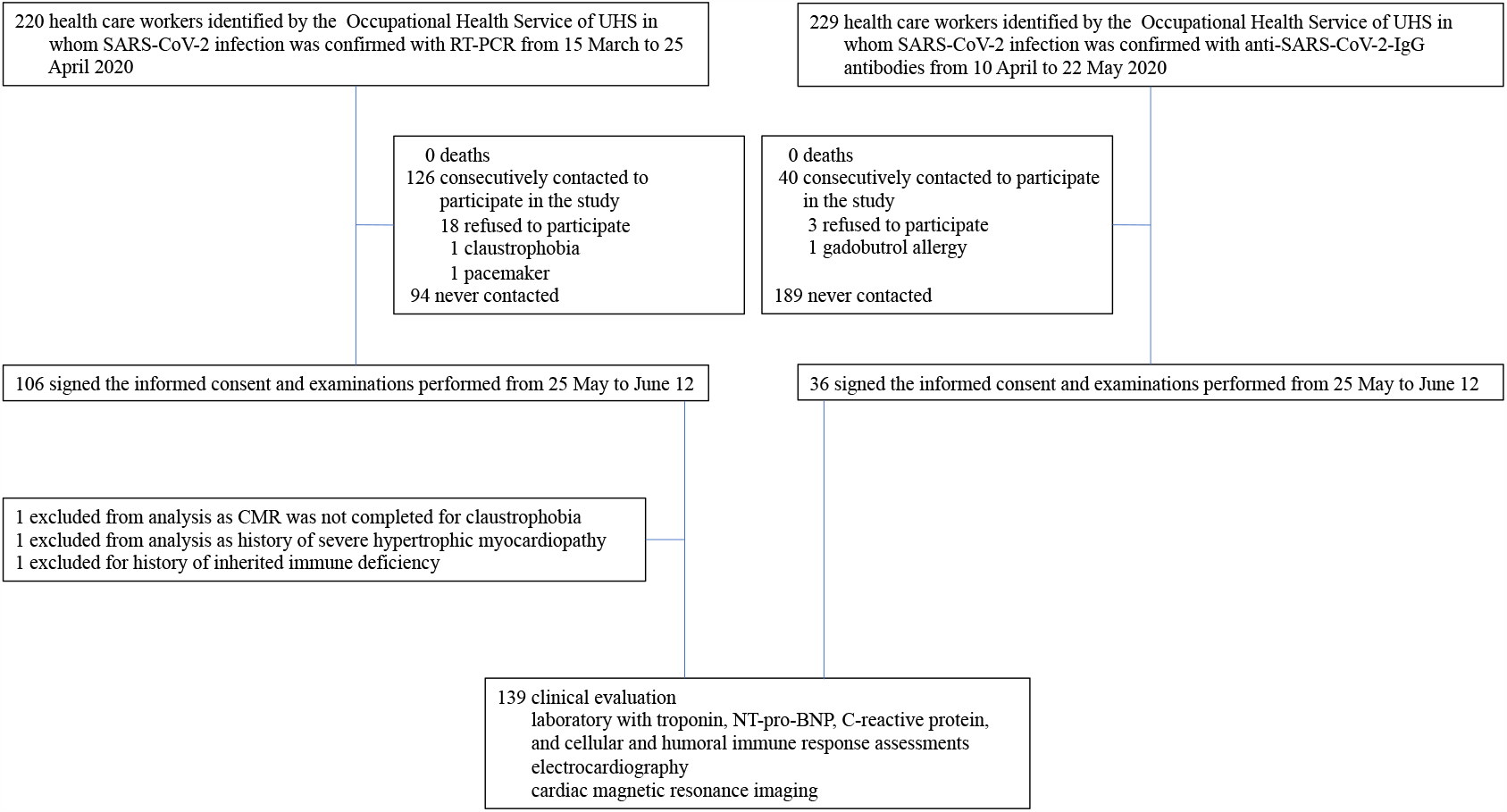
Flowchart for participant selection among health-care workers. All clinical suspicions of SARS-CoV-2 infection, RT-PCR and anti-SARS-CoV-2 antibodies test management, result information, and preventive and isolation actions for Salamanca’s health-care workers have been managed through the Occupational Health Service from the University Hospital of Salamanca during the COVID-19 pandemic; making it easy to identify health-care workers candidates for the study.

All participant characteristics are shown in **table 1**. Median age was 52 years (41–57) and most were female, 100 (72%). By professional categories, 49 (35%) were nurses, 35 (25%) medical doctors, and the remaining 55 (40%) included different profiles such as auxiliary nurses and other hospital staff. A total of 67 (48%) health-care workers were infected while directly attending covid-19 hospitalization wards.

**Table 1.**
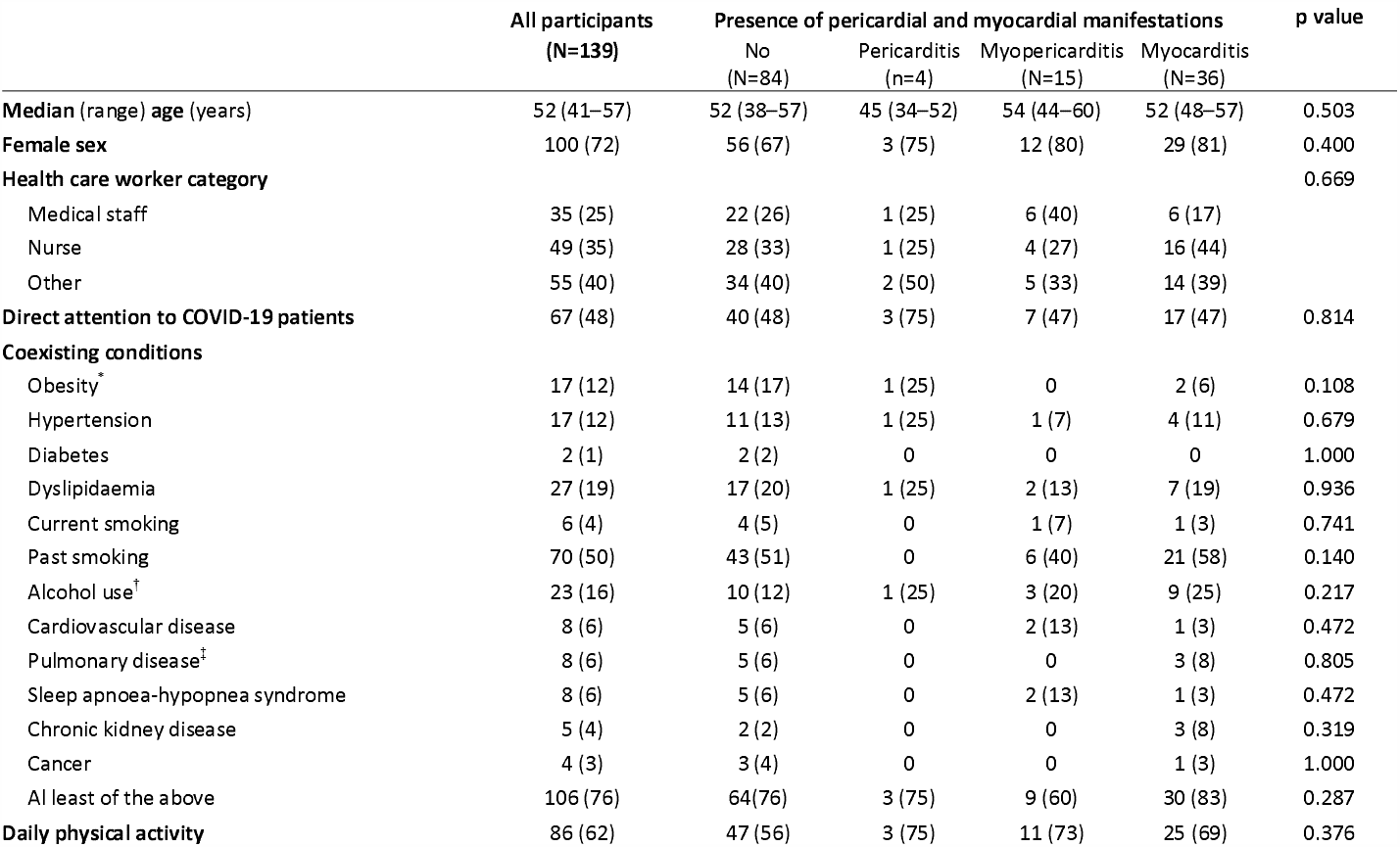
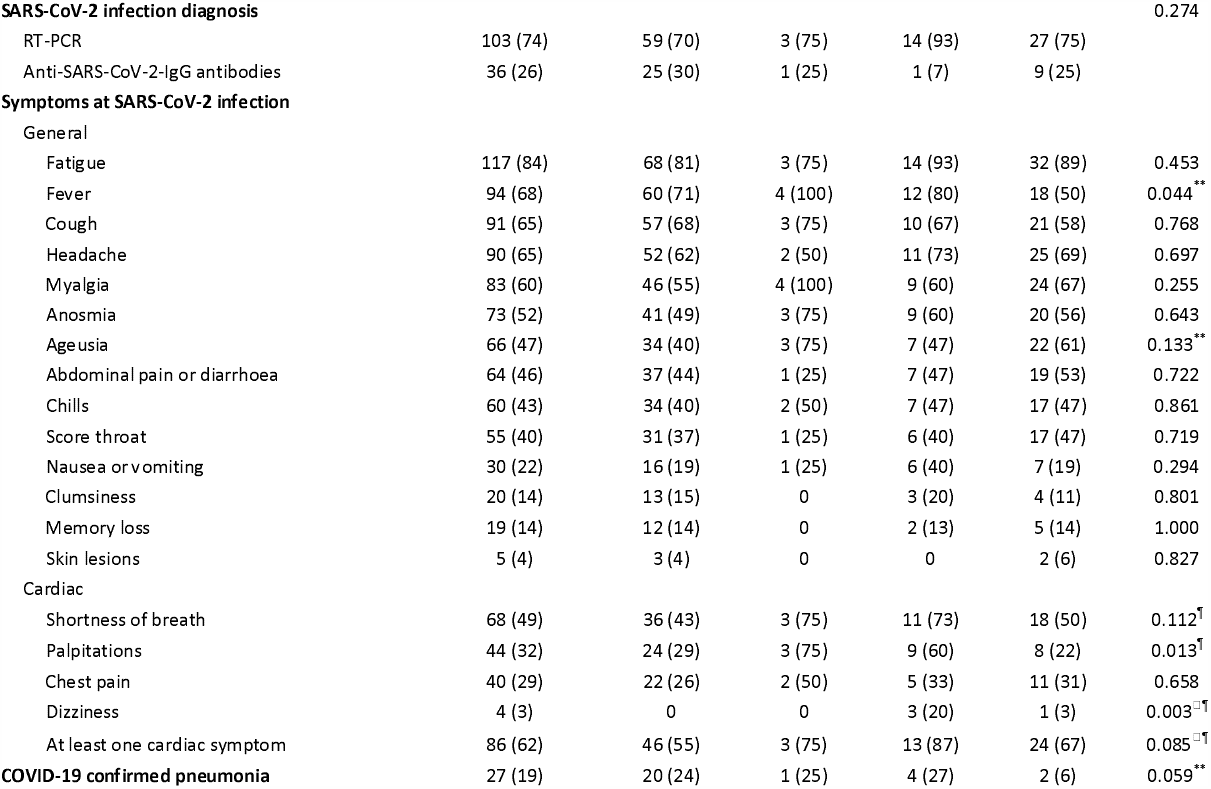
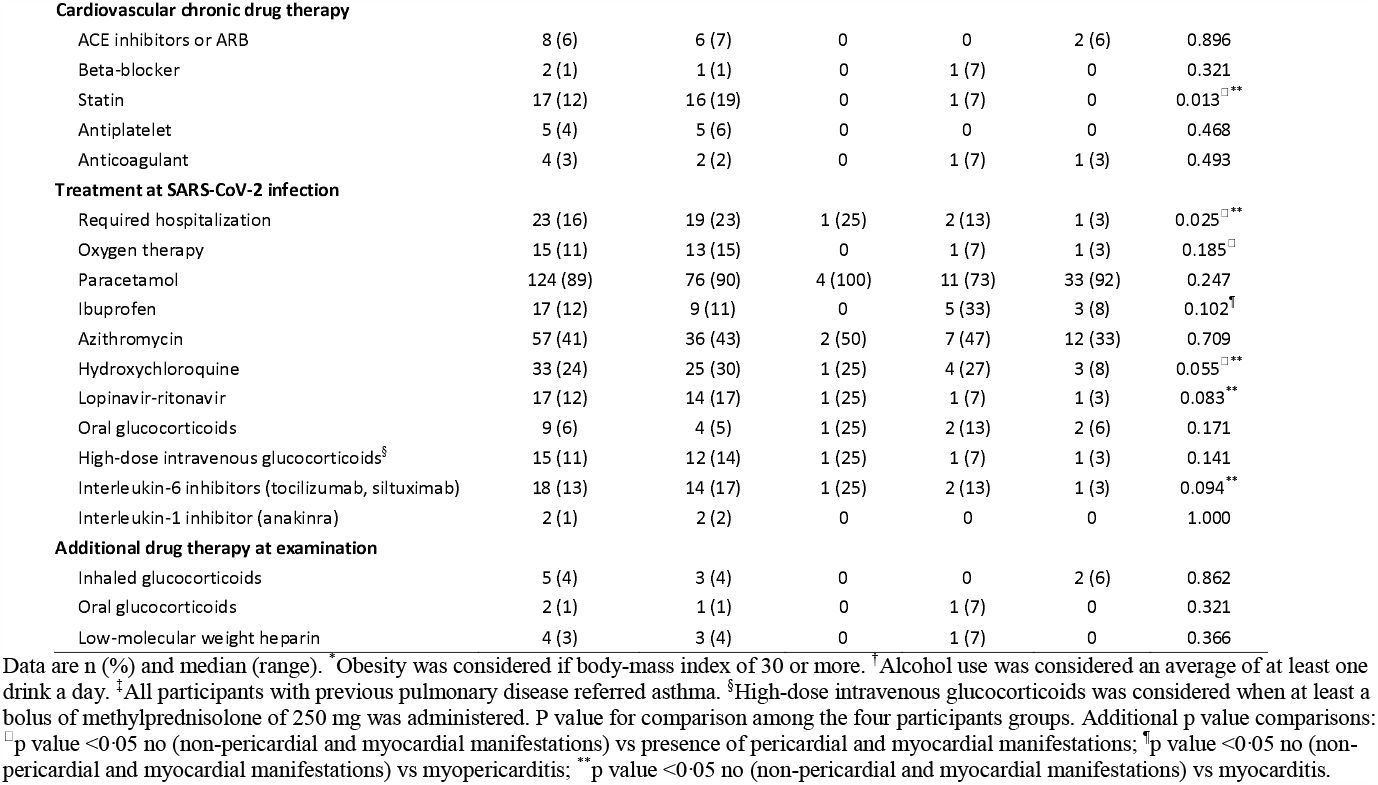
Baseline characteristics of the study cohort.

|  | All participants<br>(N=139) | Presence of pericardial and myocardial manifestations |  |  |  | p value |
| --- | --- | --- | --- | --- | --- | --- |
|  |  | No<br>(N=84) | Pericarditis<br>(n=4) | Myopericarditis<br>(N=15) | Myocarditis<br>(N=36) |  |
| **Median (range) age (years)** | 52 (41–57) | 52 (38–57) | 45 (34–52) | 54 (44–60) | 52 (48–57) | 0.503 |
| **Female sex** | 100 (72) | 56 (67) | 3 (75) | 12 (80) | 29 (81) | 0.400 |
| **Health care worker category** |  |  |  |  |  | 0.669 |
| Medical staff | 35 (25) | 22 (26) | 1 (25) | 6 (40) | 6 (17) |  |
| Nurse | 49 (35) | 28 (33) | 1 (25) | 4 (27) | 16 (44) |  |
| Other | 55 (40) | 34 (40) | 2 (50) | 5 (33) | 14 (39) |  |
| **Direct attention to COVID-19 patients** | 67 (48) | 40 (48) | 3 (75) | 7 (47) | 17 (47) | 0.814 |
| **Coexisting conditions** |  |  |  |  |  |  |
| Obesity* | 17 (12) | 14 (17) | 1 (25) | 0 | 2 (6) | 0.108 |
| Hypertension | 17 (12) | 11 (13) | 1 (25) | 1 (7) | 4 (11) | 0.679 |
| Diabetes | 2 (1) | 2 (2) | 0 | 0 | 0 | 1.000 |
| Dyslipidaemia | 27 (19) | 17 (20) | 1 (25) | 2 (13) | 7 (19) | 0.936 |
| Current smoking | 6 (4) | 4 (5) | 0 | 1 (7) | 1 (3) | 0.741 |
| Past smoking | 70 (50) | 43 (51) | 0 | 6 (40) | 21 (58) | 0.140 |
| Alcohol use<sup>†</sup> | 23 (16) | 10 (12) | 1 (25) | 3 (20) | 9 (25) | 0.217 |
| Cardiovascular disease | 8 (6) | 5 (6) | 0 | 2 (13) | 1 (3) | 0.472 |
| Pulmonary disease<sup>‡</sup> | 8 (6) | 5 (6) | 0 | 0 | 3 (8) | 0.805 |
| Sleep apnoea-hypopnea syndrome | 8 (6) | 5 (6) | 0 | 2 (13) | 1 (3) | 0.472 |
| Chronic kidney disease | 5 (4) | 2 (2) | 0 | 0 | 3 (8) | 0.319 |
| Cancer | 4 (3) | 3 (4) | 0 | 0 | 1 (3) | 1.000 |
| At least of the above | 106 (76) | 64 (76) | 3 (75) | 9 (60) | 30 (83) | 0.287 |
| **Daily physical activity** | 86 (62) | 47 (56) | 3 (75) | 11 (73) | 25 (69) | 0.376 |
|  |  | No<br>(N=84) | Pericarditis<br>(n=4) | Myopericarditis<br>(N=15) | Myocarditis<br>(N=36) |  |
| **SARS-CoV-2 infection diagnosis** |  |  |  |  |  | 0.274 |
| RT-PCR | 103 (74) | 59 (70) | 3 (75) | 14 (93) | 27 (75) |  |
| Anti-SARS-CoV-2-IgG antibodies | 36 (26) | 25 (30) | 1 (25) | 1 (7) | 9 (25) |  |
| **Symptoms at SARS-CoV-2 infection** |  |  |  |  |  |  |
| General |  |  |  |  |  |  |
| Fatigue | 117 (84) | 68 (81) | 3 (75) | 14 (93) | 32 (89) | 0.453 |
| Fever | 94 (68) | 60 (71) | 4 (100) | 12 (80) | 18 (50) | 0.044** |
| Cough | 91 (65) | 57 (68) | 3 (75) | 10 (67) | 21 (58) | 0.768 |
| Headache | 90 (65) | 52 (62) | 2 (50) | 11 (73) | 25 (69) | 0.697 |
| Myalgia | 83 (60) | 46 (55) | 4 (100) | 9 (60) | 24 (67) | 0.255 |
| Anosmia | 73 (52) | 41 (49) | 3 (75) | 9 (60) | 20 (56) | 0.643 |
| Ageusia | 66 (47) | 34 (40) | 3 (75) | 7 (47) | 22 (61) | 0.133** |
| Abdominal pain or diarrhoea | 64 (46) | 37 (44) | 1 (25) | 7 (47) | 19 (53) | 0.722 |
| Chills | 60 (43) | 34 (40) | 2 (50) | 7 (47) | 17 (47) | 0.861 |
| Sore throat | 55 (40) | 31 (37) | 1 (25) | 6 (40) | 17 (47) | 0.719 |
| Nausea or vomiting | 30 (22) | 16 (19) | 1 (25) | 6 (40) | 7 (19) | 0.294 |
| Clumsiness | 20 (14) | 13 (15) | 0 | 3 (20) | 4 (11) | 0.801 |
| Memory loss | 19 (14) | 12 (14) | 0 | 2 (13) | 5 (14) | 1.000 |
| Skin lesions | 5 (4) | 3 (4) | 0 | 0 | 2 (6) | 0.827 |
| Cardiac |  |  |  |  |  |  |
| Shortness of breath | 68 (49) | 36 (43) | 3 (75) | 11 (73) | 18 (50) | 0.112<sup>§</sup> |
| Palpitations | 44 (32) | 24 (29) | 3 (75) | 9 (60) | 8 (22) | 0.013<sup>§</sup> |
| Chest pain | 40 (29) | 22 (26) | 2 (50) | 5 (33) | 11 (31) | 0.658 |
| Dizziness | 4 (3) | 0 | 0 | 3 (20) | 1 (3) | 0.003<sup>□§</sup> |
| At least one cardiac symptom | 86 (62) | 46 (55) | 3 (75) | 13 (87) | 24 (67) | 0.085<sup>□§</sup> |
| **COVID-19 confirmed pneumonia** | 27 (19) | 20 (24) | 1 (25) | 4 (27) | 2 (6) | 0.059** |
|  |  | No<br>(N=84) | Pericarditis<br>(n=4) | Myopericarditis<br>(N=15) | Myocarditis<br>(N=36) |  |
| Cardiovascular chronic drug therapy |  |  |  |  |  |  |
| ACE inhibitors or ARB | 8 (6) | 6 (7) | 0 | 0 | 2 (6) | 0.896 |
| Beta-blocker | 2 (1) | 1 (1) | 0 | 1 (7) | 0 | 0.321 |
| Statin | 17 (12) | 16 (19) | 0 | 1 (7) | 0 | 0.013<sup>□**</sup> |
| Antiplatelet | 5 (4) | 5 (6) | 0 | 0 | 0 | 0.468 |
| Anticoagulant | 4 (3) | 2 (2) | 0 | 1 (7) | 1 (3) | 0.493 |
| Treatment at SARS-CoV-2 infection |  |  |  |  |  |  |
| Required hospitalization | 23 (16) | 19 (23) | 1 (25) | 2 (13) | 1 (3) | 0.025<sup>□**</sup> |
| Oxygen therapy | 15 (11) | 13 (15) | 0 | 1 (7) | 1 (3) | 0.185<sup>□</sup> |
| Paracetamol | 124 (89) | 76 (90) | 4 (100) | 11 (73) | 33 (92) | 0.247 |
| Ibuprofen | 17 (12) | 9 (11) | 0 | 5 (33) | 3 (8) | 0.102<sup>†</sup> |
| Azithromycin | 57 (41) | 36 (43) | 2 (50) | 7 (47) | 12 (33) | 0.709 |
| Hydroxychloroquine | 33 (24) | 25 (30) | 1 (25) | 4 (27) | 3 (8) | 0.055<sup>□**</sup> |
| Lopinavir-ritonavir | 17 (12) | 14 (17) | 1 (25) | 1 (7) | 1 (3) | 0.083<sup>**</sup> |
| Oral glucocorticoids | 9 (6) | 4 (5) | 1 (25) | 2 (13) | 2 (6) | 0.171 |
| High-dose intravenous glucocorticoids<sup>§</sup> | 15 (11) | 12 (14) | 1 (25) | 1 (7) | 1 (3) | 0.141 |
| Interleukin-6 inhibitors (tocilizumab, siltuximab) | 18 (13) | 14 (17) | 1 (25) | 2 (13) | 1 (3) | 0.094<sup>**</sup> |
| Interleukin-1 inhibitor (anakinra) | 2 (1) | 2 (2) | 0 | 0 | 0 | 1.000 |
| Additional drug therapy at examination |  |  |  |  |  |  |
| Inhaled glucocorticoids | 5 (4) | 3 (4) | 0 | 0 | 2 (6) | 0.862 |
| Oral glucocorticoids | 2 (1) | 1 (1) | 0 | 1 (7) | 0 | 0.321 |
| Low-molecular weight heparin | 4 (3) | 3 (4) | 0 | 1 (7) | 0 | 0.366 |

Data are n (%) and median (range). \*Obesity was considered if body-mass index of 30 or more. †Alcohol use was considered an average of at least one drink a day. ‡All participants with previous pulmonary disease referred asthma. §High-dose intravenous glucocorticoids was considered when at least a bolus of methylprednisolone of 250 mg was administered. P value for comparison among the four participants groups. Additional p value comparisons: <sup>□</sup>p value <0.05 no (non-pericardial and myocardial manifestations) vs presence of pericardial and myocardial manifestations; <sup>†</sup>p value <0.05 no (non-pericardial and myocardial manifestations) vs myopericarditis; <sup>\*\*</sup>p value <0.05 no (non-pericardial and myocardial manifestations) vs myocarditis.

Among the overall study population, 106 (76%) had at least one comorbidity and 8 (6%) health-care workers presented a history of cardiovascular disease; one with chronic ischemia with stent revascularization, three with paroxysmal atrial fibrillation, two with intranodal supraventricular tachycardias treated with ablation, and two with an episode of acute pericarditis several years before.

Most (137 [98%]) health-care workers experienced a viral prodrome at SARS-CoV-2 infection. Fatigue was reported by 117 (84%) participants, fever by 94 (68%), cough by 91 (65%), headache by 90 (65%) and myalgia by 83 (60%). Cardiac symptoms with shortness of breath, chest pain, palpitations or dizziness were reported by 86 (62%) participants.

A total of 27 (19%) health-care workers were previously diagnosed with covid-19 pneumonia and 23 (16%) required hospitalization. Overall, the drug therapy aimed at ameliorating the disease was heterogeneous: hydroxychloroquine was given in 33 (24%) cases, lopinavir-ritonavir in 17 (12%), oral glucocorticoids in 9 (6%), high-dose intravenous bolus of methylprednisolone in 15 (11%), and interleukin inhibitors in 18 (13%).

### Symptoms, electrocardiographic, biochemical, and cardiac magnetic resonance profiling

The study examinations (**table 2**) were performed 10·4 (9·3–11·0) weeks after symptoms of infection began; 9·4 (8·1–10·0) weeks after the positive test for RT-PCR participants and 4·4 (3·6–5·0) weeks for participants diagnosed through antibodies testing.

**Table 2.**
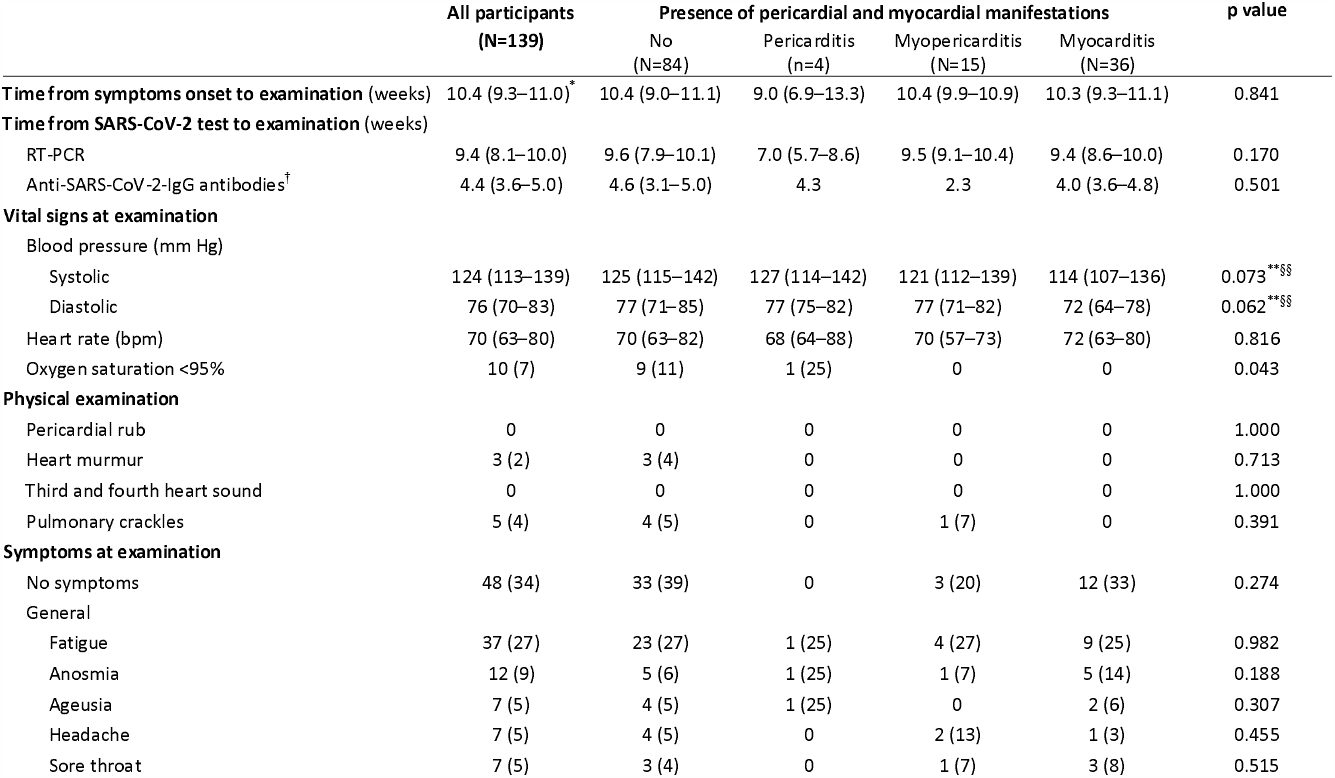
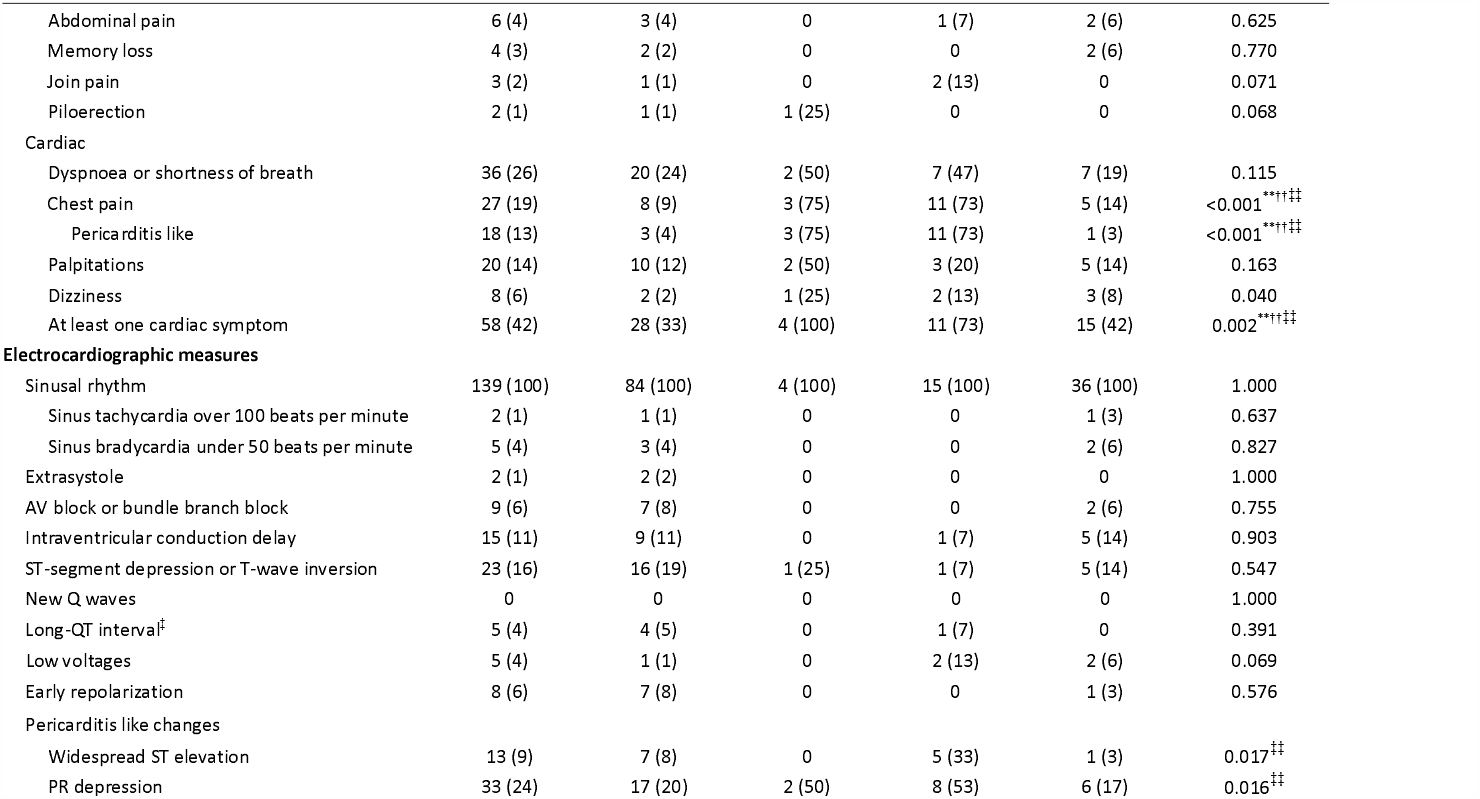
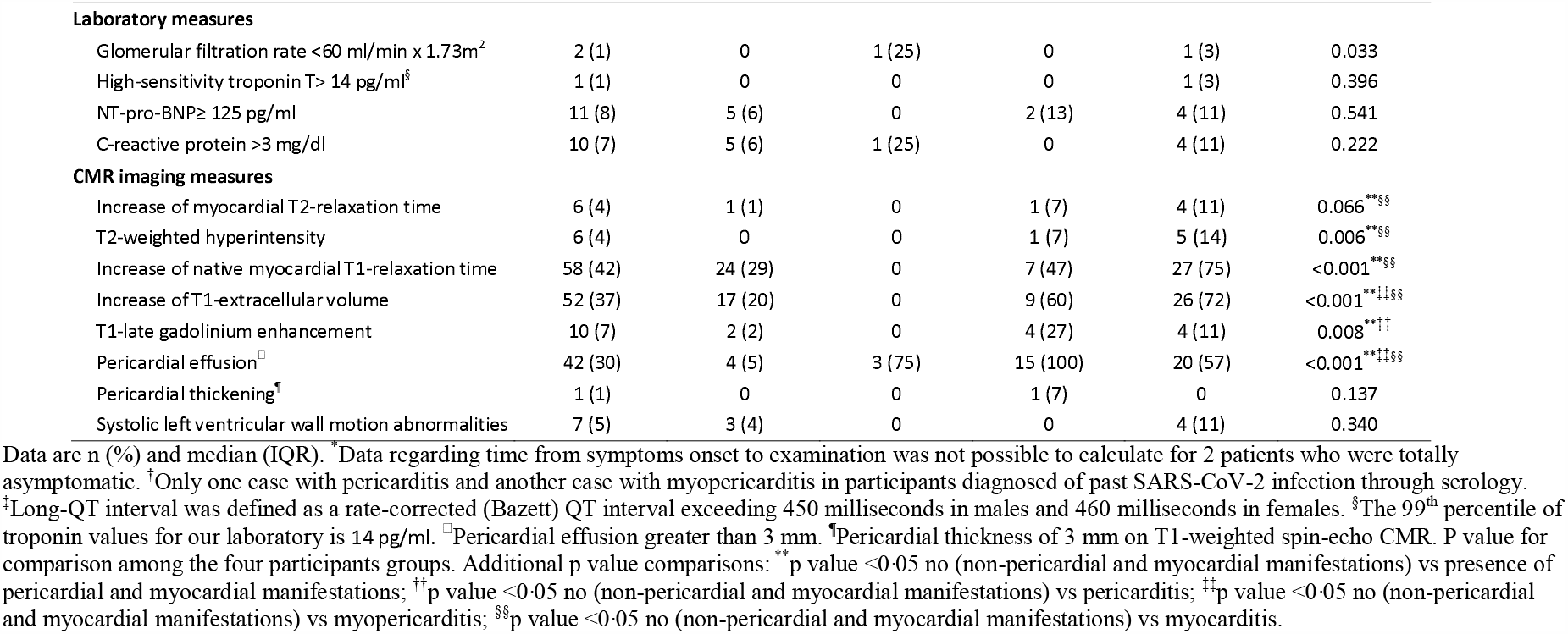
Study measures at examination.

|  | All participants<br>(N=139) | Presence of pericardial and myocardial manifestations |  |  |  | p value |
| --- | --- | --- | --- | --- | --- | --- |
|  |  | No<br>(N=84) | Pericarditis<br>(n=4) | Myopericarditis<br>(N=15) | Myocarditis<br>(N=36) |  |
| **Time from symptoms onset to examination (weeks)** | 10.4 (9.3–11.0)* | 10.4 (9.0–11.1) | 9.0 (6.9–13.3) | 10.4 (9.9–10.9) | 10.3 (9.3–11.1) | 0.841 |
| **Time from SARS-CoV-2 test to examination (weeks)** |  |  |  |  |  |  |
| RT-PCR | 9.4 (8.1–10.0) | 9.6 (7.9–10.1) | 7.0 (5.7–8.6) | 9.5 (9.1–10.4) | 9.4 (8.6–10.0) | 0.170 |
| Anti-SARS-CoV-2-IgG antibodies<sup>†</sup> | 4.4 (3.6–5.0) | 4.6 (3.1–5.0) | 4.3 | 2.3 | 4.0 (3.6–4.8) | 0.501 |
| **Vital signs at examination** |  |  |  |  |  |  |
| Blood pressure (mm Hg) |  |  |  |  |  |  |
| Systolic | 124 (113–139) | 125 (115–142) | 127 (114–142) | 121 (112–139) | 114 (107–136) | 0.073***§ |
| Diastolic | 76 (70–83) | 77 (71–85) | 77 (75–82) | 77 (71–82) | 72 (64–78) | 0.062***§ |
| Heart rate (bpm) | 70 (63–80) | 70 (63–82) | 68 (64–88) | 70 (57–73) | 72 (63–80) | 0.816 |
| Oxygen saturation <95% | 10 (7) | 9 (11) | 1 (25) | 0 | 0 | 0.043 |
| **Physical examination** |  |  |  |  |  |  |
| Pericardial rub | 0 | 0 | 0 | 0 | 0 | 1.000 |
| Heart murmur | 3 (2) | 3 (4) | 0 | 0 | 0 | 0.713 |
| Third and fourth heart sound | 0 | 0 | 0 | 0 | 0 | 1.000 |
| Pulmonary crackles | 5 (4) | 4 (5) | 0 | 1 (7) | 0 | 0.391 |
| **Symptoms at examination** |  |  |  |  |  |  |
| No symptoms | 48 (34) | 33 (39) | 0 | 3 (20) | 12 (33) | 0.274 |
| General |  |  |  |  |  |  |
| Fatigue | 37 (27) | 23 (27) | 1 (25) | 4 (27) | 9 (25) | 0.982 |
| Anosmia | 12 (9) | 5 (6) | 1 (25) | 1 (7) | 5 (14) | 0.188 |
| Ageusia | 7 (5) | 4 (5) | 1 (25) | 0 | 2 (6) | 0.307 |
| Headache | 7 (5) | 4 (5) | 0 | 2 (13) | 1 (3) | 0.455 |
| Sore throat | 7 (5) | 3 (4) | 0 | 1 (7) | 3 (8) | 0.515 |
|  |  | No<br>(N=84) | Pericarditis<br>(n=4) | Myopericarditis<br>(N=15) | Myocarditis<br>(N=36) |  |
| Abdominal pain | 6 (4) | 3 (4) | 0 | 1 (7) | 2 (6) | 0.625 |
| Memory loss | 4 (3) | 2 (2) | 0 | 0 | 2 (6) | 0.770 |
| Join pain | 3 (2) | 1 (1) | 0 | 2 (13) | 0 | 0.071 |
| Piloerection | 2 (1) | 1 (1) | 1 (25) | 0 | 0 | 0.068 |
| **Cardiac** |  |  |  |  |  |  |
| Dyspnoea or shortness of breath | 36 (26) | 20 (24) | 2 (50) | 7 (47) | 7 (19) | 0.115 |
| Chest pain | 27 (19) | 8 (9) | 3 (75) | 11 (73) | 5 (14) | <0.001<sup>***</sup>†† |
| Pericarditis like | 18 (13) | 3 (4) | 3 (75) | 11 (73) | 1 (3) | <0.001<sup>***</sup>†† |
| Palpitations | 20 (14) | 10 (12) | 2 (50) | 3 (20) | 5 (14) | 0.163 |
| Dizziness | 8 (6) | 2 (2) | 1 (25) | 2 (13) | 3 (8) | 0.040 |
| At least one cardiac symptom | 58 (42) | 28 (33) | 4 (100) | 11 (73) | 15 (42) | 0.002<sup>***</sup>†† |
| **Electrocardiographic measures** |  |  |  |  |  |  |
| Sinusal rhythm | 139 (100) | 84 (100) | 4 (100) | 15 (100) | 36 (100) | 1.000 |
| Sinus tachycardia over 100 beats per minute | 2 (1) | 1 (1) | 0 | 0 | 1 (3) | 0.637 |
| Sinus bradycardia under 50 beats per minute | 5 (4) | 3 (4) | 0 | 0 | 2 (6) | 0.827 |
| Extrasystole | 2 (1) | 2 (2) | 0 | 0 | 0 | 1.000 |
| AV block or bundle branch block | 9 (6) | 7 (8) | 0 | 0 | 2 (6) | 0.755 |
| Intraventricular conduction delay | 15 (11) | 9 (11) | 0 | 1 (7) | 5 (14) | 0.903 |
| ST-segment depression or T-wave inversion | 23 (16) | 16 (19) | 1 (25) | 1 (7) | 5 (14) | 0.547 |
| New Q waves | 0 | 0 | 0 | 0 | 0 | 1.000 |
| Long-QT interval<sup>‡</sup> | 5 (4) | 4 (5) | 0 | 1 (7) | 0 | 0.391 |
| Low voltages | 5 (4) | 1 (1) | 0 | 2 (13) | 2 (6) | 0.069 |
| Early repolarization | 8 (6) | 7 (8) | 0 | 0 | 1 (3) | 0.576 |
| **Pericarditis like changes** |  |  |  |  |  |  |
| Widespread ST elevation | 13 (9) | 7 (8) | 0 | 5 (33) | 1 (3) | 0.017<sup>††</sup> |
| PR depression | 33 (24) | 17 (20) | 2 (50) | 8 (53) | 6 (17) | 0.016<sup>††</sup> |
|  |  | No<br>(N=84) | Pericarditis<br>(n=4) | Myopericarditis<br>(N=15) | Myocarditis<br>(N=36) |  |
| **Laboratory measures** |  |  |  |  |  |  |
| Glomerular filtration rate <60 ml/min x 1.73m<sup>2</sup> | 2 (1) | 0 | 1 (25) | 0 | 1 (3) | 0.033 |
| High-sensitivity troponin T > 14 pg/ml<sup>§</sup> | 1 (1) | 0 | 0 | 0 | 1 (3) | 0.396 |
| NT-pro-BNP ≥ 125 pg/ml | 11 (8) | 5 (6) | 0 | 2 (13) | 4 (11) | 0.541 |
| C-reactive protein >3 mg/dl | 10 (7) | 5 (6) | 1 (25) | 0 | 4 (11) | 0.222 |
| **CMR imaging measures** |  |  |  |  |  |  |
| Increase of myocardial T2-relaxation time | 6 (4) | 1 (1) | 0 | 1 (7) | 4 (11) | 0.066<sup>**§§</sup> |
| T2-weighted hyperintensity | 6 (4) | 0 | 0 | 1 (7) | 5 (14) | 0.006<sup>**§§</sup> |
| Increase of native myocardial T1-relaxation time | 58 (42) | 24 (29) | 0 | 7 (47) | 27 (75) | <0.001<sup>**§§</sup> |
| Increase of T1-extracellular volume | 52 (37) | 17 (20) | 0 | 9 (60) | 26 (72) | <0.001<sup>**††§§</sup> |
| T1-late gadolinium enhancement | 10 (7) | 2 (2) | 0 | 4 (27) | 4 (11) | 0.008<sup>**††</sup> |
| Pericardial effusion<sup>□</sup> | 42 (30) | 4 (5) | 3 (75) | 15 (100) | 20 (57) | <0.001<sup>**††§§</sup> |
| Pericardial thickening<sup>¶</sup> | 1 (1) | 0 | 0 | 1 (7) | 0 | 0.137 |
| Systolic left ventricular wall motion abnormalities | 7 (5) | 3 (4) | 0 | 0 | 4 (11) | 0.340 |

Data are n (%) and median (IQR). \*Data regarding time from symptoms onset to examination was not possible to calculate for 2 patients who were totally asymptomatic. †Only one case with pericarditis and another case with myopericarditis in participants diagnosed of past SARS-CoV-2 infection through serology. ‡Long-QT interval was defined as a rate-corrected (Bazett) QT interval exceeding 450 milliseconds in males and 460 milliseconds in females. §The 99<sup>th</sup> percentile of troponin values for our laboratory is 14 pg/ml. □ Pericardial effusion greater than 3 mm. ¶ Pericardial thickness of 3 mm on T1-weighted spin-echo CMR. P value for comparison among the four participants groups. Additional p value comparisons: \*\*p value <0.05 no (non-pericardial and myocardial manifestations) vs presence of pericardial and myocardial manifestations; ††p value <0.05 no (non-pericardial and myocardial manifestations) vs pericarditis; ‡‡p value <0.05 no (non-pericardial and myocardial manifestations) vs myopericarditis; §§p value <0.05 no (non-pericardial and myocardial manifestations) vs myocarditis.

All participants presented vital and exploratory signs of hemodynamic stability at examination. 91 (65%) health-care workers still presented symptoms with fatigue being reported in 37 (27%) cases, and those cardiac-related, such as shortness of breath in 36 (26%) cases, chest pain in 27(19%), or palpitations in 20 (14%).

Of the 139 electrocardiograms, electrocardiographic abnormalities were reported in 69 (50%) cases. Minnesota codes abnormalities were observed in 49 (35%) participants (supplementary table A), QT interval prolongation was observed in 5 (4%), and early repolarization in 8 (6%). A total of 33 (24%) electrocardiograms met criteria for pericarditis-like changes (supplementary figure A).

Cardiac-specific and inflammatory biomarkers were within the normal range in most participants. Above the normal range, high-sensitivity troponin T concentration (≥14 pg. per millilitre) was increased in 1 (1%) participant, NT-pro-BNP (≥125 pg. per millilitre) in 11 (8%) and C-reactive protein (≥3 mg. per decilitre) in 10 (7%).

CMR abnormalities were observed in 104 (75%) cases (complete data from CMR measures are in the supplementary tables B, C and D). 6 (4%) participants presented increased of myocardial T2-relaxation time, 6 (4%) oedema on T2-weighted images, 58 (42%) increased native myocardial T1-relaxation time, 52 (37%) increase of T1-extracellular volume, 10 (7%) T1-late gadolinium enhancement, 42 (30%) pericardial effusion, 1 (1%) a pericardial thickness of 3 mm and 7 (5%) systolic left ventricular wall motion abnormalities, global or regional (supplementary Figure B).

### Pericarditis and myocarditis prevalence

A total of 55 (40%) participants fulfilled criteria for either pericarditis or myocarditis. Among them, 19 (14%) fulfilled criteria for pericarditis, 51 (37%) criteria for myocarditis, and 15 (11%) fulfilled criteria for both pericarditis and myocarditis.

Of the 19 participants fulfilling criteria for pericarditis, 15 (79%) presented two classical criteria and 4 (21%) three criteria. Of the 51 participants fulfilling criteria for myocarditis, 22 (43%) presented a combination of at least two T2 or T1-based LLC main criteria and 29 (57%) presented a combination of only one T2 or T1-based LLC main criterion with at least one additional LLC supportive criterion. Additional descriptions of criteria combinations are provided in **figure 2**.

**Figure 2:**
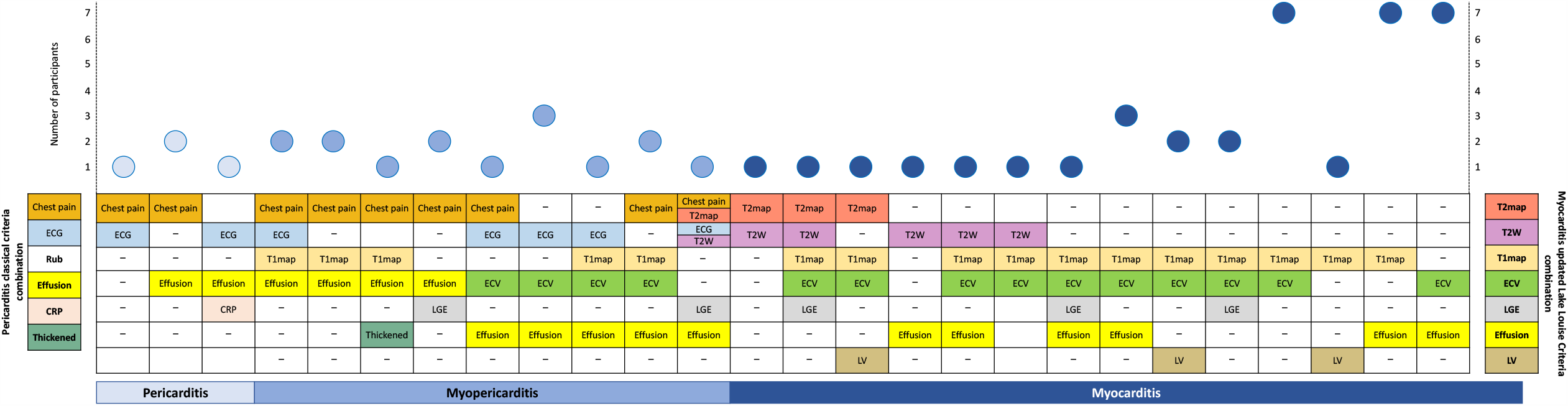
Pericarditis, myopericarditis and myocarditis criteria combinations. Description of pericarditis clinical criterions and updated Lake-Louise-Criteria for myocarditis in participants diagnosed with pericarditis, myopericarditis or myocarditis. CRP= C-reactive protein; ECG= electrocardiogram; ECV= increase of T1-extracellular volume; Effusion= pericardial effusion assessed at cardiac magnetic resonance; LGE= T1-late gadolinium enhancement; LV= systolic left ventricular wall motion abnormalities; T1map= increase of native myocardial T1-relaxation time; T2 map= increase of myocardial T2-relaxation time; T2W= increase of T2-weighted hyperintensity; Thickened= pericardial thickness greater or equal to 3 mm.

Clinical isolated pericarditis was then diagnosed in 4 (3%) cases, myopericarditis in 15 (11%) and isolated myocarditis in 36 (26%). Baseline and examination characteristics for each diagnostic group are detailed in **tables 1** and **2**.

A higher percentage of participants without pericardial or myocardial manifestations than participants with pericarditis, myopericarditis and myocarditis were on chronic drug therapy with statins (16 [19%] vs. 1 [2%]; p=0.003), were previously hospitalized for covid-19 (19 [23%] vs. 4 [7%]; p=0.020), and received drug therapy aimed at ameliorating the disease: hydroxychloroquine (25 [30%] vs. 8 [14%]; p=0.043), lopinavir-ritonavir (14 [17%] vs. 3 [5%]; p=0·064) and interleukin inhibitors or high-doses of intravenous glucocorticoids (16 [19%] vs. 4 [7%]; p=0.082). A higher percentage of participants with pericarditis, myopericarditis or myocarditis than participants without these manifestations presented cardiac symptoms at SARS-CoV-2 infection (40 [73%] vs. 46 [55%]; p=0.049) and at study examination (30 [54%] vs. 28 [33%]; p=0.015).

### Participants with infection confirmed through anti-SARS-CoV-2-IgG detection

Among the participants diagnosed with past infection through anti-SARS-CoV-2-IgG detection (data for this group, compared to RT-PCR participants, are shown in the appendix [supplementary table E), 28 (78%) were previously tested negative by RT-PCR after developing mild SARS-CoV-2 symptoms and 8 (22%) were never RT-PCR tested. A lower percentage of participants diagnosed through positive serology still presented symptoms at examination compared to RT-PCR participants (18 [50%] vs. 73 [71%]; p=0.027); nonetheless, the prevalence of pericarditis, myopericarditis or myocarditis was high in both groups (**figure 3**).

**Figure 3:**
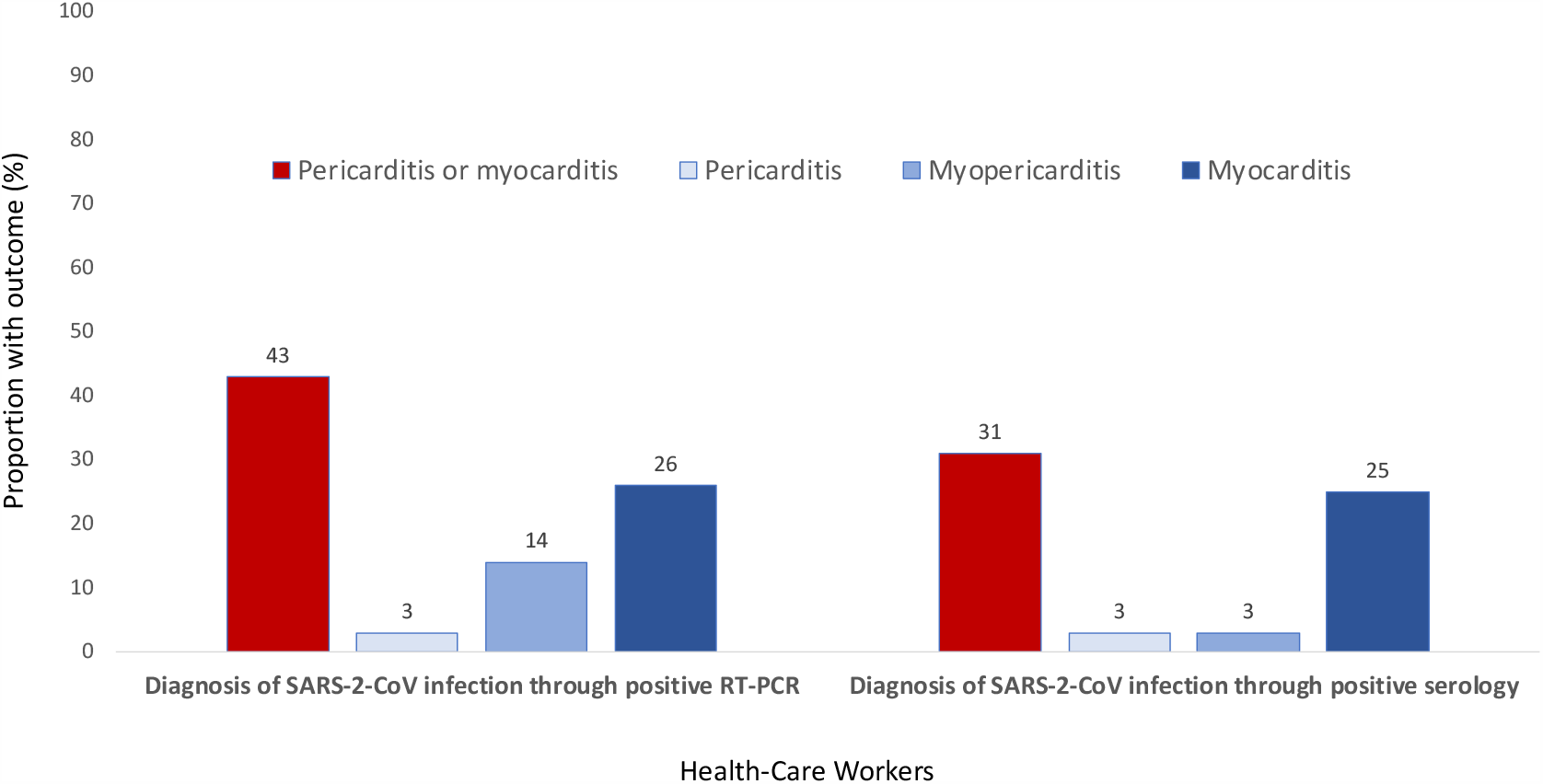
Pericarditis and myocarditis prevalence by SARS-CoV-2 infection laboratory diagnosis. Prevalence of pericarditis or myocarditis, pericarditis, myopericarditis and myocarditis in participants with SARS-CoV-2 infection diagnosed through RT-PCR or through serology.

### Altered immune cell and humoral profiles in blood

.Most study participants (101 [73%]) displayed altered cell counts in blood for at least one major immune cell population as illustrated in **figure 4A-B**, and described in more detail in **tables 3** and **4**. Most frequent alterations consisted of eosinopenia (38 [27%] cases; p<0.001), and increased CD4^-^CD8^-/lo^ Tαβ (cytotoxic) T-cell (24 [17%] cases; p<0.001); and, to a less extent also B-cell counts (16 [12%] cases; p<0.001). In addition, compared to age-matched healthy donors, participants had higher median counts in blood of basophils (38 vs. 47 cells/μL, respectively; p<0.013), total monocytes (317 vs 405 cells/μL; p<0.001) at the expense of classical (FcεRI^-^) monocytes (290 vs 386 cells/μL; p<0 001), total lymphocytes (1675 vs 2221 cells/μL; p<0.001), including total T-lymphocytes (1246 vs 1652 cells/μL; p<0.001), TCD4 (716 vs 1017 cells/μL; p<0.001), TCD8 (407 vs 507 cells/μL; p<0·001) and B lymphocytes (154 vs 208 cells/μL; p<0.001); in contrast, participants showed decreased median numbers of circulating blood myeloid-derived suppressor cells (5 vs 2 cells/μL; p<0.001, immature CD16^-^ CD62L^-^ neutrophils (5 vs 2 cells/μL; p<0 001), intermediate monocytes (20 vs 14 cells/μL; p=0·001) and immunomodulatory FcεRI^+^CD62L^+^(46 vs 12; p=0.038) and FcεRI^+^CD62L^-^ (7 vs 2; p=0.001) monocytes, plasmacytoid dendritic cells (8 vs 6 cells/μL; p=0.020), NK-cells (260 vs 213 cells/μL; p=0.007) and circulating plasma cells (2 vs 0·8 cells/μL; p<0.001).

**Table 3.** Distribution of subsets of myeloid immune cells in blood.

|  | Healthy donors<br>(N=463) | All participants<br>(N=139) | p value | Presence of pericardial and myocardial manifestations |  |  |  | p value |
| --- | --- | --- | --- | --- | --- | --- | --- | --- |
|  |  |  |  | No<br>(N=84) | Pericarditis<br>(n=4) | Myopericarditis<br>(N=15) | Myocarditis<br>(N=36) |  |
| **Neutrophils** | 3723 (2969–4613) | 3430 (2633–4225) | 0.004<sup>*§</sup> | 3413 (2647–4122) | 2767 (2386–5975) | 3734 (3121–4249) | 3290 (2318–4407) | 0.650 |
| % cases ↓5<sup>th</sup>p/↑95<sup>th</sup>p | – | 18 (13) / 5 (5) | <0.001 / 0.001 | 9 (11) / 5 (6) | 0 / 0 | 1 (7) / 0 | 8 (22) / 2 (6) | 0.299 / 0.794 |
| MDSC-like Neutrophils | 5 (2–12) | 2 (1–5) | <0.001<sup>**§</sup> | 3 (1–8) | 2 (0.7–28) | 2 (0.7–3) | 2 (1–4) | 0.270 |
| Immature CD16<sup>+</sup>CD62L<sup>–</sup> | 5 (2–9) | 2 (0.8–4) | <0.001<sup>‡§</sup> | 2 (0.9–6) | 2 (0.6–23) | 0.9 (0.6–2) | 2 (0.8–3) | 0.280 |
| Immature CD16<sup>+</sup>CD62L<sup>+</sup> | 0.5 (0.2–1) | 0.5 (0.3–1) | 0.920 | 0.5 (0.3–1) | 0.5 (0.2–5) | 0.3 (0.1–0.9) | 0.4 (0.2–0.8) | 0.270 |
| **Eosinophils** | 157 (101–249) | 74 (47–149) | <0.001<sup>**‡§</sup> | 82 (50–120) | 34 (31–164) | 61 (46–126) | 105 (50–179) | 0.340 |
| % cases ↓5<sup>th</sup>p/↑95<sup>th</sup>p | – | 38 (27) / 1 (0.7) | <0.001 / 0.200 | 21 (25) / 1 (1) | 3 (75) / 0 | 4 (27) / 0 | 10 (28) / 0 | 0.840 / 0.883 |
| **Basophils** | 38 (21–52) | 47 (35–62) | 0.013<sup>‡d</sup> | 48 (37–52) | 34 (13–55) | 34 (29–56) | 51 (38–64) | 0.210 |
| % cases ↓5<sup>th</sup>p/↑95<sup>th</sup>p | – | 4 (2) / 5 (3) | 0.001 / 0.001 | 1 (1) / 3 (3) | 1 (25) / 1 (25) | 1 (6) / 1 (6) | 1 (2) / 0 | 0.842 / 0.672 |
| **Monocytes** | 317 (245–433) | 405 (328–523) | <0.001<sup>*</sup> | 421 (346–528) | 376 (351–785) | 385 (297–442) | 368 (292–557) | 0.330<sup>□¶</sup> |
| % cases ↓5<sup>th</sup>p/↑95<sup>th</sup>p | – | 7 (5) / 6 (4) | <0.001/ <0.001 | 4 (5) / 3 (4) | 0/ 1 (25) | 3 (20) / 0 | 0 / 2 (6) | 0.185 / 0.650 |
| Classical monocytes | 290 (240–383) | 386 (308–492) | <0.001<sup>*</sup> | 423 (356–500) | 382 (319–669) | 376 (308–518) | 341 (272–409) | 0.059<sup>□¶</sup> |
| CD62L<sup>+</sup>FcγRI<sup>+</sup> | 46 (7–70) | 12 (3–56) | 0.038<sup>‡§</sup> | 18 (5–73) | 4 (1–15) | 2 (0.7–6) | 11 (3–34) | 0.020<sup>¶</sup> |
| CD62L<sup>+</sup>FcγRI<sup>–</sup> | 205 (132–269) | 230 (156–304) | 0.170<sup>*</sup> | 245 (160–330) | 263 (248–580) | 189 (54–293) | 196 (156–252) | 0.057<sup>¶</sup> |
| CD62L<sup>–</sup>FcγRI<sup>+</sup> | 7 (2–33) | 2 (0.3–7) | 0.001<sup>**‡§</sup> | 2 (0.3–9) | 0.4 (0.1–4) | 0.5 (0.1–7) | 2 (0.3–7) | 0.740 |
| CD62L<sup>–</sup>FcγRI<sup>–</sup> | 101 (52–258) | 39 (18–77) | <0.001<sup>**‡§</sup> | 35 (16–77) | 23 (13–36) | 49 (19–100) | 47 (28–72) | 0.560 |
| Intermediate monocytes | 20 (15–29) | 14 (9–20) | 0.001<sup>**‡§</sup> | 15 (10–21) | 9 (4–14) | 13 (5–17) | 13 (9–18) | 0.150 |
| Non-classical monocytes | 49 (36–62) | 42 (27–63) | 0.170 | 43 (27–63) | 36 (13–51) | 45 (11–73) | 39 (27–58) | 0.730 |
| **Dendritic cells** | 29 (20–35) | 28 (20–35) | 0.620 | 29 (21–35) | 28 (22–31) | 29 (18–36) | 23 (17–35) | 0.420 |
| % cases ↓5<sup>th</sup>p/↑95<sup>th</sup>p | – | 14 (10) / 5 (3) | <0.001 / 0.001 | 7 (8) / 5 (5) | 1 (25) / 0 | 6 (40) / 0 | 0 / 0 | 0.556 / 0.343 |
| Myeloid dendritic cells | 18 (14–26) | 19 (14–26) | 0.600 | 19 (14–25) | 20 (14–26) | 21 (15–29) | 17 (13–28) | 0.800 |
| CD1c<sup>+</sup> m dendritic cells | 17 (14–25) | 19 (14–25) | 0.570 | 19 (14–25) | 19 (13–26) | 20 (14–27) | 16 (13–24) | 0.790 |
| CD141<sup>+</sup> m dendritic cells | 1 (0.6–1) | 0.9 (0.6–1) | 0.640 | 0.9 (0.6–1) | 0.6 (0.5–0.7) | 0.8 (0.7–1) | 0.6 (0.5–1) | 0.054 |
| Plasmacytoid dendritic cells | 8 (6–11) | 6 (4–9) | 0.020<sup>§</sup> | 7 (5–10) | 7 (2–12) | 6 (4–8) | 5 (4–8) | 0.063 |
| Axl<sup>+</sup> dendritic cells | 0.3 (0.2–0.4) | 0.3 (0.2–0.5) | 0.560 | 0.3 (0.2–0.5) | 0.3 (0.2–0.4) | 0.3 (0.2–0.5) | 0.2 (0.1–0.3) | 0.330 |

Data are median (IQR) of cells/ μL of blood or n (%) of cases lower than 5th percentile ( $\downarrow 5^{\text{th}}$ p) and higher than 95th percentile ( $\uparrow 95^{\text{th}}$ p) for each cell subset in age-matched healthy donors. Left p value for comparisons between healthy donors vs all participants. Right p value for comparison among the four participants groups. Additional p value comparisons: \*p value <0.05 healthy donors vs no (non-pericardial and myocardial manifestations); $^{\dagger}$ p value <0.05 healthy donors vs pericarditis; $^{\ddagger}$ p value <0.05 healthy donors vs myopericarditis; $^{\S}$ p value <0.05 healthy donors vs myocarditis; $^{\square}$ p value <0.05 no (non-pericardial and myocardial manifestations) vs myocarditis. $^{\dagger}$ p value <0.05 no (non-pericardial and myocardial manifestations) vs myopericarditis plus myocarditis. MDSC= myeloid-derived suppressor cells; m= myeloid.

**Table 4.**
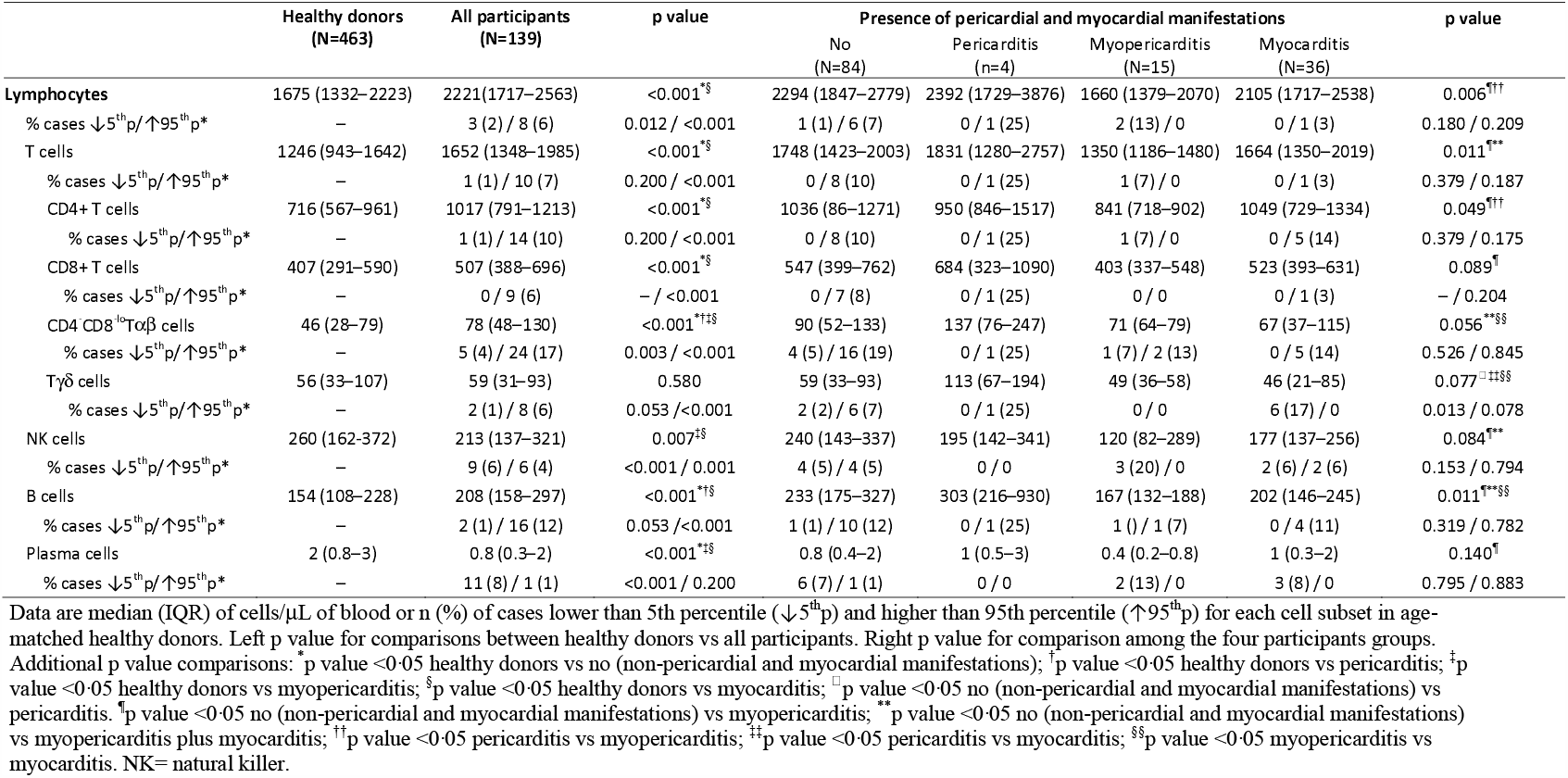
Distribution of the major subsets of lymphoid cells in blood.

**Figure 4:**
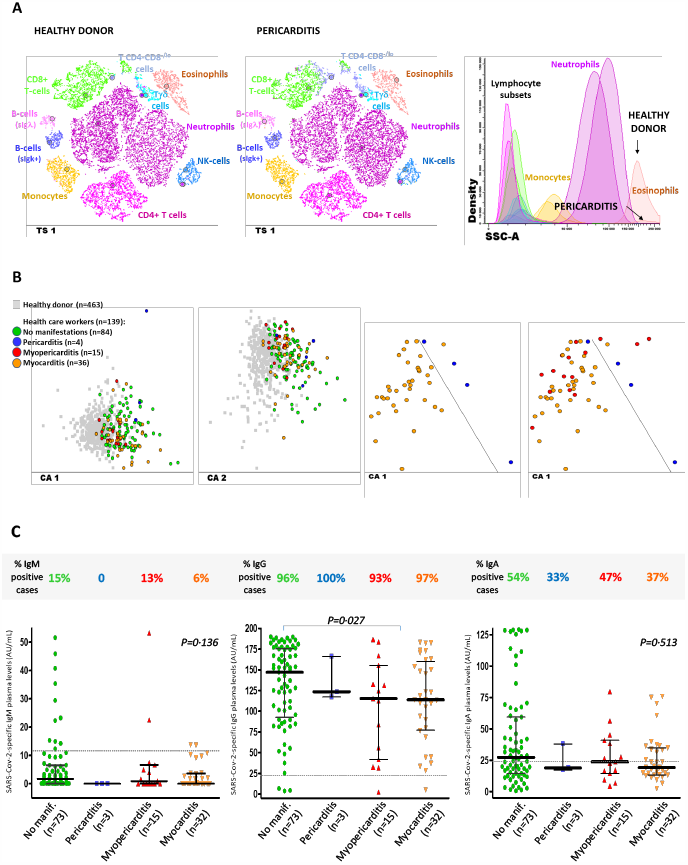
Altered immune cell profiles and antibody serum levels in the health-care workers grouped according to the presence vs absence of pericarditis, myopericarditis and myocarditis. A: t-SNE graphical representation of the distribution of the major immune cell populations in blood of a healthy donor (T-SNE plot in the left) and a participant diagnosed with pericarditis (t-SNE middle plot) showing both increased Tab+CD4-CD8-/lo T cell and decreased eosinophil counts in blood. B: 2-dimension graphical representation of multivariate canonical (linear discriminant) analysis plots showing the presence of overall different immune cell profiles in blood of the participants with past SARS-CoV-2 infection (n=139; coloured circles) vs age-matched healthy donors (n=463; grey squares) (two plots in the left); distinct immune cell profiles were also observed between participants diagnosed with pericarditis (blue circles) and myocarditis (orange circles); most myopericarditis cases (red circles) shared an altered immune cell profile in blood similar to that of myocarditis. C: Frequency and amount of anti-SARS-CoV-2 antibodies measured in plasma of 123/139 participants grouped according to the absence vs presence of pericarditis, myopericarditis and myocarditis.

Compared to healthy donors, participants with pericarditis showed the highest median counts in blood of CD4^-^CD8^-/lo^ Tαβ (cytotoxic) T-cells (46 vs 137 cells/μL; p=0.011) and also displayed (similarly to cases with myopericarditis and those with myocarditis) decreased blood counts of FcεRI^+^CD62L^+^ immunomodulatory monocytes (46 vs 4, 2 and 11, respectively; p=0.020).

Participants diagnosed with myopericarditis tended to share altered immune cell profiles with myocarditis, except for two cases with myopericarditis whose profiles more closely overlapped with those cases with pericarditis (**figure 4B**). Thus, compared to healthy donors, participants with myopericarditis and those with myocarditis specifically displayed more pronounced decreased counts in blood of plasmacytoid dendritic cells (8 vs 6 and 5 cells/μL, respectively; p<0.001), and NK-cells (260 vs 120 and 177 cells/μL; p<0.001), including the subset of cytotoxic granzyme B (Gz)^+^CD57^-^ of NK-cell, Tγδ and/or TCD8 cells (supplementary table F), and circulating plasma cells (2 vs 0·4 and 1 cells/μL; p<0.001) (supplementary table G).

Finally, participants with myopericarditis and those with myocarditis presented lower amounts of anti-SARS-CoV-2-IgG antibodies (115 [42–156] and 114 [77–160] IU/mL, respectively) compared to participants without pericardial or myocardial manifestations (147 [93–176] IU/mL; p=0.027) (**figure 4C**). Of note, among participants with myopericarditis, the above alterations were further associated with lower TCD4 Th17 lymphocytes (20 vs 33 cells/μL; p=0.023) (supplementary table H).

## Discussion

This study examined the prevalence of pericarditis and of myocarditis in a cohort of SARS-CoV-2 positive health-care workers in Salamanca, Spain. In the largest cohort of subjects with CMR imaging assessment reported so far, we demonstrate that pericardial and myocardial involvement is highly prevalent after SARS-CoV-2 infection.

We decided to carry out a study in health-care workers, as this sector has been disproportionally infected in Spain —approximately 20% of all COVID-19 cases^21^— which provided us with the opportunity to study the prevalence of pericarditis and myocarditis in SARS-CoV-2 infected cases that were confirmed by positive RT-PCR or positive serology. In addition, as the proportion of female health-care workers is high in Spain, our study does not underrepresent women who constituted more than two thirds of recruited participants. Unlike other observational studies suggesting that myocarditis may be slightly more prevalent in men than women^22-24^; men in our study presented lower prevalence of pericarditis or myocarditis than women (11 [28%] vs. 44 [44%]; p=0.122). This observation could be related, more than to gender, to the higher rate of previous hospitalization for COVID-19 pneumonia in men than women (13 [33%] vs. 10 [10%]; p=0.002) for our cohort. In general, previously hospitalized participants presented lower percentage of pericardial and myocardial injury compared to participants never hospitalized (4 [17%] vs. 51 [44%]; p=0.020). Having been hospitalized, participants received more treatment aimed at inhibiting viral replication (lopinavir-ritonavir or hydroxychloroquine) and reducing inflammation (high-dose glucocorticoids and interleukin inhibitors). The recent Recovery study (NCT04381936) has shown that low-dose dexamethasone reduces mortality in hospitalized COVID-19 patients.

Case reports of myocarditis and pericarditis have been published during the COVID-19 hospitalization phase^25^. A recent retrospective observation in 26 recovered patients with COVID-19 pneumonia presenting cardiac complaints during hospitalization, revealed the presence of myocardial oedema in 14 (54%) patients and late gadolinium enhancement in 8 (31%) patients^26^. Our results are in agreement with these findings as 89 (64%) of our participants presented myocardial injury in T2 or in T1-based CMR. High rates of myocardial damage are also observed in influenza —where elevated cardiac enzymes, electrocardiographic, echocardiographic and histologic findings have been reported in approximately one third of cases^27^.

Importantly, clinical assessment of our participants with pericarditis and myocarditis showed clinical stability without any participant presenting severe pericardial effusion, heart failure or left ventricular dysfunction (only four participants with myocarditis presented wall motion abnormalities). However, follow-up studies are necessary to determine the outcome of cardiac sequelae observed even in asymptomatic and pauci-symptomatic subjects after SARS-CoV-2 infection^28^. Thus, the participants diagnosed of past infection through serology who were more likely to be asymptomatic or mildly symptomatic, and who might better represent the cases detected in population-wide seroprevalence studies^11,12^, presented a similar prevalence of pericardial and myocardial manifestations to RT-PCR positive participants (11 [31%] vs 44 [43%]; p=0.238).

At present, there is much interest in the long-term sequelae of COVID-19. It is intriguing that pericarditis, myopericarditis or myocarditis were observed that long after SARS-CoV-2 infection (over 10 weeks after initial viral prodrome at infection) and also in some presently asymptomatic subjects (15 [11%] of the study population; one every three-final pericarditis, myopericarditis or myocarditis diagnosis). These long-term manifestations may be due to an inadequate innate and adaptative immune response. In recent months, important advances have been achieved in the understanding of the immunology of COVID-19^10^.

However, there is still very limited data on the longer-term immunological consequences of past SARS-CoV-2 infection, and no study has specifically focused in the settings of pericarditis and myocarditis. Herein, in-depth investigation of the distribution of major and minor populations of immune cells in blood showed a high frequency of overall altered immune profiles. Several of the immune cell alterations identified mimic abnormalities reported during active infection, including decreased eosinophil, NK-cell, and (plasmacytoid) dendritic cell counts^29,30^. In contrast, other alterations are described here for the first time and have not been reported during the acute phase of the infection. These new alterations include abnormally high numbers of monocytes (i.e. recently produced classical monocytes) and lymphocytes, such as T-cells, particularly CD4^-^CD8^-/lo^, TCD4 cells and TCD8 cells, and B-lymphocytes together with decreased counts of myeloid-derived suppressor cells, immature (CD16-CD62L-) neutrophils, both immunomodulatory FcεRI^+^CD62L^+^ and intermediate (i.e. CD14+CD16+) monocytes and circulating plasma cells. Altogether, these findings may partially reflect (the normal and an altered) immune recovery after SARS-CoV-2 infection. Thus, while persistence of eosinopenia and expansion of CD4^-^CD8^-/lo^ T-cells in blood were detected across all groups of participants suggesting it is part of the immune recovery after COVID-19; NK-cells were specifically decreased among participants with myopericarditis and myocarditis in association with (persistent) tissue damage. Nonetheless, the altered immune profiles in participants without pericardial and myocardial manifestations, could also be due to persistence of tissue damage and recovery in other organs affected by SARS-CoV-2, such as the lung, more than just reflecting normal immune recovery. Anyhow, more detailed analysis of the altered immune profiles among the different groups of participants showed that those with myopericarditis or myocarditis had closer to normal lymphocyte counts, but reduced numbers in blood of circulating eosinophils, plasmacytoid dendritic cells and particularly, NK-cells. Such unique profile mimics what has been described recently during the acute phase of SARS-CoV-2 infection, suggesting an ongoing cytotoxic response with increased tissue migration or death by apoptosis of specific subsets of cytotoxic cells. In line with this hypothesis, we found here lower numbers of granzymeB+CD57-memory and effector TCD8-, Tγδ- and/or NK-cells among participants with myopericarditis or myocarditis. Of note, these participants with myocardial injury also showed particularly lower counts in blood of (recently produced) plasma cells, together with lower anti-SARS-CoV-2-IgG plasma levels. These findings suggest that a less pronounced (potentially insufficient) or a delayed humoral response may occur in these subjects, which may lead to decreased neutralization, opsonization and/or clearance of the virus locally at the peri-myocardium; local viral persistence would favour an increased tissue-homing (or early death) of eosinophils, immunomodulatory and intermediate monocytes, in addition to cytotoxic (effector) cells. Altogether, the above findings suggest that an inadequate (potentially delayed) immune response might happen in a substantial fraction of patients with past SARS-CoV-2 infection, with differently altered profiles in subjects who present myopericarditis or myocarditis.

### Limitations of the study

The study analysis was limited to health-care workers in Salamanca and therefore may have limited external generalizability to other non-health-care settings. However, the strength of this study is the addition of non-hospitalized participants and also the inclusion of participants diagnosed of past SARS-CoV-2 infection through serology, who also had a high prevalence of pericarditis and myocarditis. Seropositive participants, although less symptomatic than RT-PCR participants, presented mild symptoms in almost all cases; unfortunately, we cannot draw conclusions regarding the prevalence of pericarditis and myocarditis in the completely asymptomatic general population. Finally, the study relied solely on descriptive observations and cannot provide any conclusion on the benefit of antiviral and anti-inflammatory treatments during the acute phase of infection or whether the prevalence of pericarditis and myocarditis after SARS-CoV-2 infection is higher in women than men.

## Conclusions

This study shows that pericarditis and myocarditis are frequent long after SARS-CoV-2 infection and also in some presently asymptomatic subjects; in addition, we provide herein evidence for an altered immune cell distribution in blood which affects cells involved in both the innate (e.g. eosinophils and monocytes) and the adaptative cellular (e.g. CD4^-^CD8^-/lo^Tαβ^+^, TCD4 and TCD8 T-cells) and humoral (e.g. plasma cells and anti-Sars-CoV-2 IgG levels) immune responses; nonetheless, a direct link between such altered immune profiles and the presence of pericardial and myocardial injury still needs to be established. These observations will probably apply to the general population infected, and may indicate that cardiac sequelae might occur late. Although all study participants presented clinical stability and non-severe cardiac complications, prospective monitoring will be necessary to address the future clinical consequences of these findings.

## Data Availability

The deindentification data in this study will be shared upon request and approval will be designated by a data access committee. The data access committee comprises four authors and there is no restriction to data access.

## Abbreviations

CMR: cardiac magnetic resonance
CRP: C-reactive protein
ECG: electrocardiogram
ECV: extracellular volume
FcεRI: high affinity IgE receptor I
GZ: granzyme B
IMM: immunemonitoring
LGE: late gadolinium enhancement
LLC: Lake-Louise-Criteria
LV: left ventricle
LST: lymphocyte screening tube
RT-PCR: reverse-transcriptase-polymerase-chain-reaction
SARS-CoV-2: severe acute respiratory syndrome-coronavirus 2
Th: T-helper cells
t-SNE: t-distributed stochastic neighbour embedding
T1: map native myocardial T1-relaxation time
T2: map myocardial T2-relaxation time T2W
T2: -weighted hyperinstensity

## Acknowledgments

We thank all health-care workers participating in this study, to all the personnel of the University Hospital of Salamanca for their extraordinary work in their fight against this pandemic, and all other participating institutions. We also would like to thank Prof Dr Jacques JM van Dongen and both his group (Leiden University Medical Center) and the EuroFlow Consortia for their contribution in the design, construction and validation of the EuroFlow LST and IMM tubes used herein.

## Authors contribution

RE, MBP, JA, JLBG, AO, PLS conceived and designed the study; RE, MB-P, AO, PLS contributed to the literature research; RE, SM, IT contributed with clinical evaluation of the participants; SM, CS-P, IT, EDP, LMA contributed with clinical data collection; MBP, AMG contributed to data analysis (cardiac magnetic resonance); EV, DGC contributed to data analysis (ECG); APP, ATV, OPE, BFH performed immunophenotypic and serology experiments and collected data; JA, BFH, GOA, QL, RF, AO contributed to data analysis and interpretation (Immunophenotypic and serology); RE, MBP, AMG, EV, AO, PLS, contributed to clinical data analysis and interpretation; MBP, JA, JLBG, AO, PLS contributed to writing of the report.

